# Adaptive nutrition intervention stabilizes serum phosphorus levels in hemodialysis patients: a multi-center decentralized clinical trial using real-world data

**DOI:** 10.1101/2023.02.19.23286157

**Authors:** Moon Kyung Chung, Do Hyoung Kim, Ji In Park, Sunhwa Lee, Hayne Cho Park, Kyungmin Kim, Young Sun Kang, Kangji Ko, Jieun Kim, Hoseok Koo, Jin Joo Cha, Young Eun Kwon, Ju Han Kim

## Abstract

**Objective:** This study aims to evaluate the effect of an adaptive nutritional and educational intervention for hemodialysis patients in a routine care setting, using real-world data from electronic health records.

**Methods:** Decentralized clinical trial of seven hemodialysis facilities recruited patients under hemodialysis for over 3 months (N=153) for an 8-week adaptive intervention protocol and divided them into four groups: (1) control (2) education intervention (3) meal intervention (4) education and meal interventions. Educational contents are digitally delivered via mobile phones and pre-made meals tailored on laboratory findings via home delivery. Changes in serum electrolytes and malnutrition inflammation score (MIS) are analyzed.

**Results:** Meal intervention statistically significantly stabilized serum phosphorus level (β = −0.81) at week 8 with increased likelihood of being in normal range (Odds ratio = 1.21). Meal and education group showed better nutritional status (MIS=3.97) than the control group (MIS=4.57) at week 8 (adjusted p<0.05). No significant changes were observed in serum potassium level, depression, and self-efficacy.

**Conclusion:** It is demonstrated that an adaptive education and meal interventions in a real-world care setting may benefit hemodialysis patients’ serum phosphorus control and nutritional status, without negative effect on depression levels or self-efficacy.

## Introduction

Hyperkalemia and hyperphosphatemia are associated with poor prognosis in hemodialysis (HD) patients. Excess electrolytes are removed by HD but should be preemptively controlled through diet to reduce complication risk from elevated interdialytic levels. HD patients are prone to malnutrition due to nutrient loss during HD, dietary restriction, decreased appetite, and poor physical activity [1]. The renal diet is one of the most complex regimens with numerous restrictions on fruits, vegetables, nuts, legumes, whole grains, seasonings, and fluids [2]. Soaking and boiling removes potassium [3] but requires extra preparation time. Thus, patients find difficulty following dietary recommendations [4, 5], reporting more challenges in execution of meal preparation over knowledge or information [6, 7].

On one hand, recent research on the renal diet suggests liberating restrictions and tailoring to the patient’s status, to improve dietary adherence and quality of life [8, 9]. Tailored interventions improve engagement and have a small significant effect in improving health behavior and outcomes [10-13]. However, existing methods heavily rely on healthcare professionals. Computerized-tailoring methods are becoming prevalent, but not yet widely utilized in CKD [14, 15]. On the other hand, pre-made meals can help patients who are novices or environmentally constrained. Medically tailored meals for chronic disease have shown to improve nutritional status, biomarkers, and healthcare costs [16, 17]. However, meal studies for CKD have been mostly focused on sodium, protein, or specific diet and supplements [18-20].

We designed an adaptive nutritional and educational intervention protocol for HD patients with mobile education and home-delivered meals. This study is unique in that it (1) aims to tailor to each patient’s serum potassium and phosphorus levels while maintaining a good nutritional status, (2) attempts to develop an intervention method in a real-world setting using electronic health record (EHR), and (3) adopts virtual elements of decentralized clinical trials [21].

## Methods

### 2.1 Study design and patients

The study team has previously developed a decentralized personal health record platform comprised of (1) an iPad application for physicians that provide multi-center clinical data interoperability and (2) a smartphone application (App) for patients, connected to the platform in a peer-to-peer network on Ethereum blockchain that supports decentralized data privacy [22, 23]. An algorithm is designed, with guidance from nephrologists and dietitians, to generate meal delivery orders and tailored education materials within the platform. It is executed by a virtual intermediary program, utilizing real-world clinical data from routine care; no additional review from healthcare experts or lab work is needed during the trial.

Patients who are under HD for more than 3 months, over 18 years old, and not diagnosed with cancer within 5 years; are recruited from seven dialysis facilities connected to the platform. All participants are encouraged to install the App, which provides access to their clinical records and educational materials. Groups are randomized using a permuted block method with a block size of 4, stratified according to site. Randomization is executed independently from App usage, but no blinding procedures are in place. Baseline data is obtained preferably 2 weeks before the trial initiation date. If recruitment is late, the intervention initiates on subsequent Wednesdays, to match the meal production schedule. Initiation dates ranged from October 13, 2021, to November 3, 2021. Figure 1 shows a CONSORT diagram of the enrollment process.

**Figure 1.**
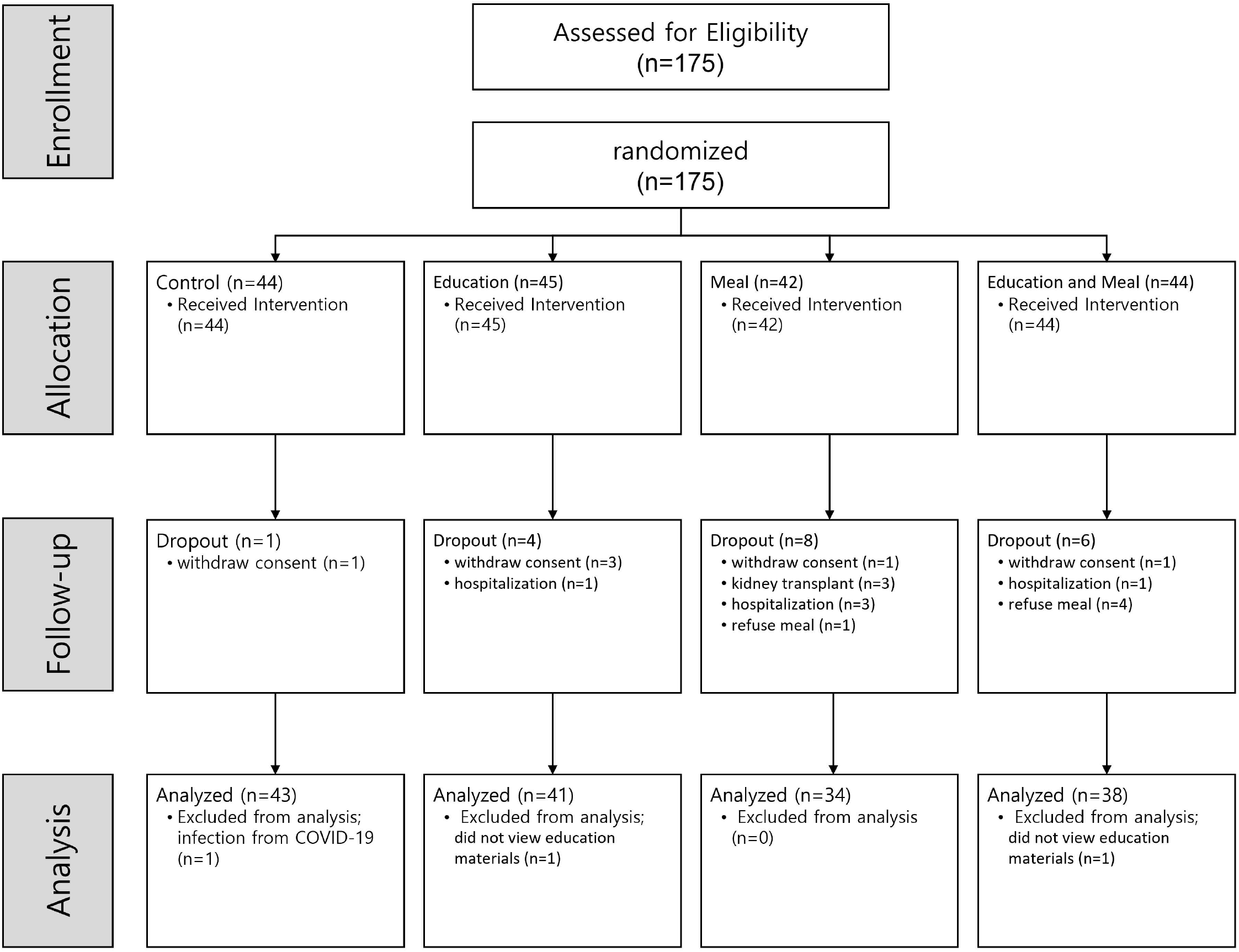
CONSORT flow diagram.

### 2.3 Intervention

Education materials should be provided via the App. However, due to limitations in development time and low App adoption rate, it was rerouted to be provided via Short Message Service (SMS) links for this study. Materials consist of 5 leaflets and 30 videos, categorized by content (Supplementary Table 1). Materials on basic nutrition, eating out, and protein are provided to all patients receiving education. Protein materials are customized based on the patient’s weight, including recommendation on daily protein intake and example meal plans. Malnutrition materials are provided based on baseline Malnutrition Inflammation Score (MIS). Potassium, phosphate, and sodium materials are provided based on the patient’s blood chemistry, prescription of binders, and interdialytic weight, respectively. Values are checked weekly for new updates. All materials are sent only once, until depletion.

Pre-made frozen meals, selected from commercially available options, are delivered to the participant’s homes. Each meal is composed of white rice, a protein dish, and three side dishes; packaged in a box. Three or four meals are delivered twice weekly with instructions to replace one meal daily. A description of nutrient composition was added mid-trial to address patients’ concerns about excess potassium. Meals are provided in two types: (A) low sodium, and (B) low sodium, potassium, and phosphate. Type B is provided if the patient’s most recent blood test results are over the threshold or if the patient is prescribed with binders. Type A is provided if none of the conditions are met. Nutrient content standards are determined by dividing the daily recommended intake for HD patients by three (sodium under 660mg, potassium under 660mg, and phosphate under 330mg). Actual nutrient content of the meals is calculated based on raw ingredients, making sure to not exceed the standards set forth. Since minerals are lost during cooking, this ensures that the nutrient content is equal to or less than the standard. Manufacture and delivery of meals are executed by a private enterprise who retails tailored meals for CKD patients.

An overview of the algorithm and delivery schedule is in Figure 2.

**Figure 2.**
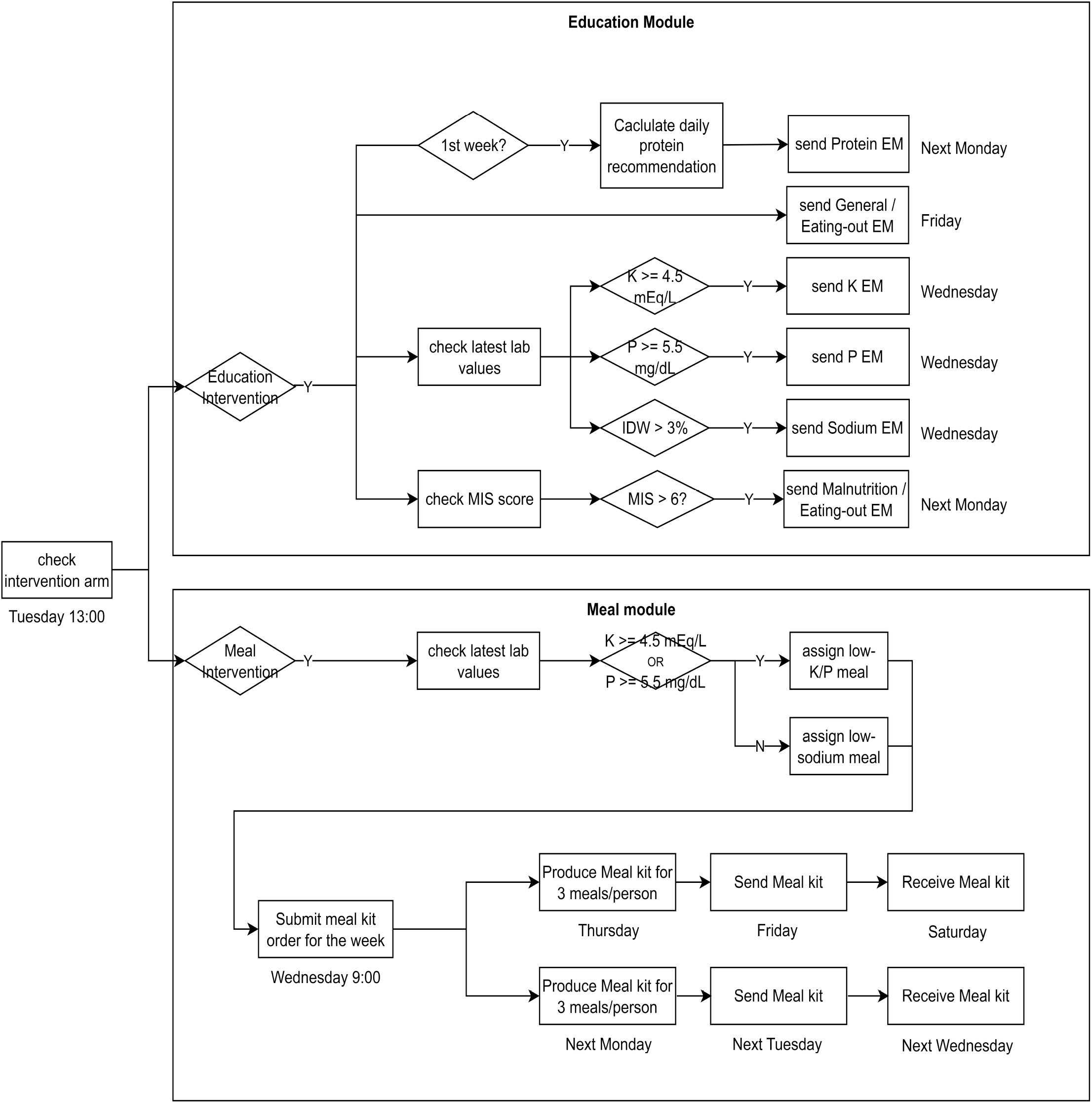
Intervention Workflow. The process was repeated weekly, starting from Tuesday until the next Monday, for a total of 8 weeks. EM: Education Materials, K: serum potassium, IDW: interdialytic weight gain, MIS: malnutrition inflammation score, P: serum phosphorus.

### 2.4 Measurements

Anthropometric measurements, vital signs, and blood test results are obtained from EHR. Since HD patients get monthly blood test in routine care, data is available at baseline (week 0), midpoint (week 4) and endpoint (week 8). Surveys consist of 4 existing tools and 2 designed for the study. Existing tools are MIS, Korean-Beck Depression Inventory (K-BDI), Short Form 36 (SF-36) [24], and Self-Efficacy survey [25]; collected at baseline and endpoint. For patients assigned with meals, meal compliance is measured by a weekly 4-point scale survey on the amount of provided meal consumed (0 : none, 1 : <1/3, 2 : 1/3∼2/3, 3 : ≥2/3); delivery failures are scored as 0. For patients assigned with education, education satisfaction is measured by a weekly 5-point Likert scale survey on whether the patient found the materials helpful.

### 2.5 Statistical Analysis

Baseline values are expressed as mean ± standard deviation for continuous variables, and frequency (percentage, %) for categorical variables. Differences in baseline characteristics are assessed using One-way Analysis of Variance (ANOVA) for continuous variables and Fisher’s Exact Test for categorical variables.

Main outcomes are serum potassium, serum phosphorus, and MIS; and secondary outcomes are K-BDI and Self-Efficacy. Significance of intervention effect is assessed with 3-way ANCOVA, using meal, education and time as factors and baseline value as covariate. For significant terms, a post-hoc t-test is conducted with Bonferroni adjustment. Effect sizes at weeks 4 and 8 are estimated using linear regression, adjusting for baseline values.

The algorithm targets to normalize, rather than invariably reduce, serum levels. Therefore, outcomes are also assessed as dichotomized variables at normal range and intervention thresholds. Normal ranges are defined as ≥3.5 and <5.5 mEq/L for serum potassium, ≥2.5 and < 6.5 mg/dL for serum phosphorus, and <6 for MIS. Intervention thresholds are ≥4.5 mEq/L for serum potassium and ≥5.5 mEq/L for serum phosphorus. Differences between groups are assessed with Fisher’s Exact Test. Likelihood of having normal serum values at weeks 4 and 8 are estimated using logistic regression, adjusting for baseline values. Exponent of regression coefficients are calculated to determine the likelihood in odds ratio.

All statistical analyses are performed using R version 4.0.2 (R Foundation for Statistical Computing; http://www.r-project.org/). Statistical significance is set at p < 0.05.

## Results

### 3.1 Baseline characteristics

Out of the 175 participants enrolled, 19 dropped out due to consent withdrawal (n=6), hospitalization (n=5), kidney transplant (n=3), and refusal of meal intervention (n=5). Three patients were excluded from analysis after intervention, due to inability to collect endpoint measurements (n=1) or having refused to open all education materials (n=2). As a result, 153 patients were included in the final analysis (Figure 1). There were no significant differences between the groups in baseline characteristics (Table 1) or App usage rates (Supplementary Table 2).

**Table 1.**
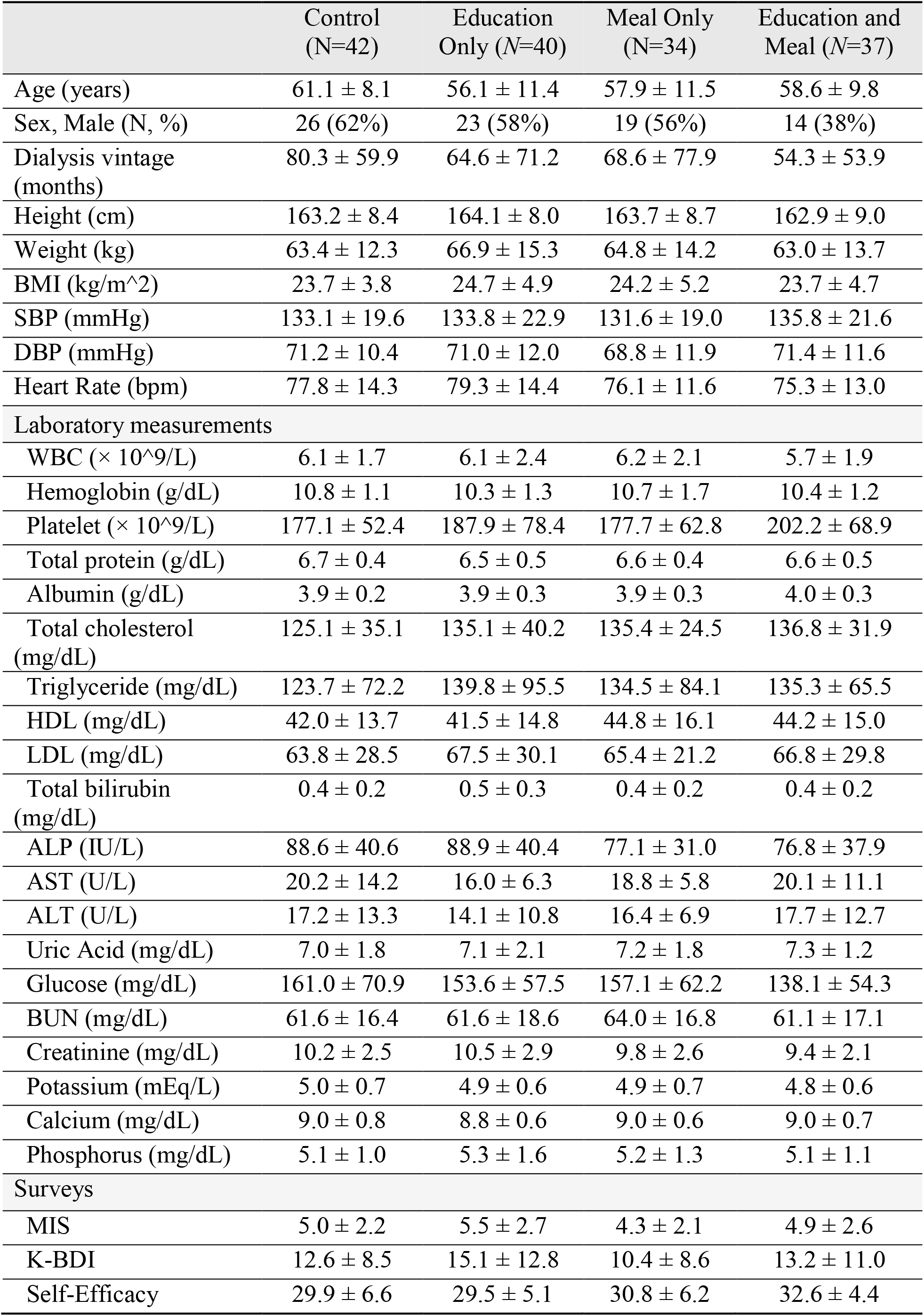
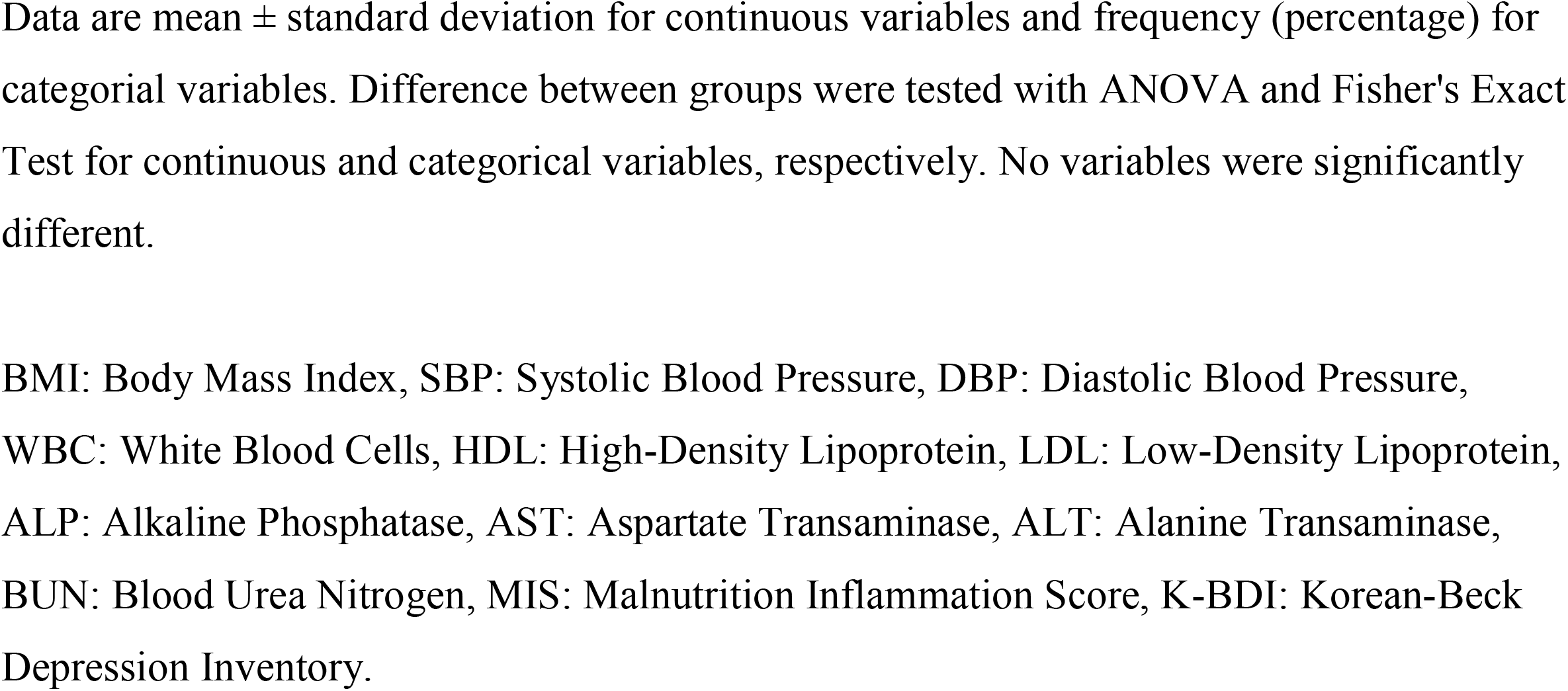
Baseline Clinical Characteristics.

|  | Control<br>(N=42) | Education<br>Only (N=40) | Meal Only<br>(N=34) | Education and<br>Meal (N=37) |
| --- | --- | --- | --- | --- |
| Age (years) | 61.1 ± 8.1 | 56.1 ± 11.4 | 57.9 ± 11.5 | 58.6 ± 9.8 |
| Sex, Male (N, %) | 26 (62%) | 23 (58%) | 19 (56%) | 14 (38%) |
| Dialysis vintage<br>(months) | 80.3 ± 59.9 | 64.6 ± 71.2 | 68.6 ± 77.9 | 54.3 ± 53.9 |
| Height (cm) | 163.2 ± 8.4 | 164.1 ± 8.0 | 163.7 ± 8.7 | 162.9 ± 9.0 |
| Weight (kg) | 63.4 ± 12.3 | 66.9 ± 15.3 | 64.8 ± 14.2 | 63.0 ± 13.7 |
| BMI (kg/m<sup>2</sup>) | 23.7 ± 3.8 | 24.7 ± 4.9 | 24.2 ± 5.2 | 23.7 ± 4.7 |
| SBP (mmHg) | 133.1 ± 19.6 | 133.8 ± 22.9 | 131.6 ± 19.0 | 135.8 ± 21.6 |
| DBP (mmHg) | 71.2 ± 10.4 | 71.0 ± 12.0 | 68.8 ± 11.9 | 71.4 ± 11.6 |
| Heart Rate (bpm) | 77.8 ± 14.3 | 79.3 ± 14.4 | 76.1 ± 11.6 | 75.3 ± 13.0 |
| Laboratory measurements |  |  |  |  |
| WBC (× 10<sup>9</sup>/L) | 6.1 ± 1.7 | 6.1 ± 2.4 | 6.2 ± 2.1 | 5.7 ± 1.9 |
| Hemoglobin (g/dL) | 10.8 ± 1.1 | 10.3 ± 1.3 | 10.7 ± 1.7 | 10.4 ± 1.2 |
| Platelet (× 10<sup>9</sup>/L) | 177.1 ± 52.4 | 187.9 ± 78.4 | 177.7 ± 62.8 | 202.2 ± 68.9 |
| Total protein (g/dL) | 6.7 ± 0.4 | 6.5 ± 0.5 | 6.6 ± 0.4 | 6.6 ± 0.5 |
| Albumin (g/dL) | 3.9 ± 0.2 | 3.9 ± 0.3 | 3.9 ± 0.3 | 4.0 ± 0.3 |
| Total cholesterol<br>(mg/dL) | 125.1 ± 35.1 | 135.1 ± 40.2 | 135.4 ± 24.5 | 136.8 ± 31.9 |
| Triglyceride (mg/dL) | 123.7 ± 72.2 | 139.8 ± 95.5 | 134.5 ± 84.1 | 135.3 ± 65.5 |
| HDL (mg/dL) | 42.0 ± 13.7 | 41.5 ± 14.8 | 44.8 ± 16.1 | 44.2 ± 15.0 |
| LDL (mg/dL) | 63.8 ± 28.5 | 67.5 ± 30.1 | 65.4 ± 21.2 | 66.8 ± 29.8 |
| Total bilirubin<br>(mg/dL) | 0.4 ± 0.2 | 0.5 ± 0.3 | 0.4 ± 0.2 | 0.4 ± 0.2 |
| ALP (IU/L) | 88.6 ± 40.6 | 88.9 ± 40.4 | 77.1 ± 31.0 | 76.8 ± 37.9 |
| AST (U/L) | 20.2 ± 14.2 | 16.0 ± 6.3 | 18.8 ± 5.8 | 20.1 ± 11.1 |
| ALT (U/L) | 17.2 ± 13.3 | 14.1 ± 10.8 | 16.4 ± 6.9 | 17.7 ± 12.7 |
| Uric Acid (mg/dL) | 7.0 ± 1.8 | 7.1 ± 2.1 | 7.2 ± 1.8 | 7.3 ± 1.2 |
| Glucose (mg/dL) | 161.0 ± 70.9 | 153.6 ± 57.5 | 157.1 ± 62.2 | 138.1 ± 54.3 |
| BUN (mg/dL) | 61.6 ± 16.4 | 61.6 ± 18.6 | 64.0 ± 16.8 | 61.1 ± 17.1 |
| Creatinine (mg/dL) | 10.2 ± 2.5 | 10.5 ± 2.9 | 9.8 ± 2.6 | 9.4 ± 2.1 |
| Potassium (mEq/L) | 5.0 ± 0.7 | 4.9 ± 0.6 | 4.9 ± 0.7 | 4.8 ± 0.6 |
| Calcium (mg/dL) | 9.0 ± 0.8 | 8.8 ± 0.6 | 9.0 ± 0.6 | 9.0 ± 0.7 |
| Phosphorus (mg/dL) | 5.1 ± 1.0 | 5.3 ± 1.6 | 5.2 ± 1.3 | 5.1 ± 1.1 |
| Surveys |  |  |  |  |
| MIS | 5.0 ± 2.2 | 5.5 ± 2.7 | 4.3 ± 2.1 | 4.9 ± 2.6 |
| K-BDI | 12.6 ± 8.5 | 15.1 ± 12.8 | 10.4 ± 8.6 | 13.2 ± 11.0 |
| Self-Efficacy | 29.9 ± 6.6 | 29.5 ± 5.1 | 30.8 ± 6.2 | 32.6 ± 4.4 |

Data are mean ± standard deviation for continuous variables and frequency (percentage) for categorical variables. Difference between groups were tested with ANOVA and Fisher's Exact Test for continuous and categorical variables, respectively. No variables were significantly different.
BMI: Body Mass Index, SBP: Systolic Blood Pressure, DBP: Diastolic Blood Pressure, WBC: White Blood Cells, HDL: High-Density Lipoprotein, LDL: Low-Density Lipoprotein, ALP: Alkaline Phosphatase, AST: Aspartate Transaminase, ALT: Alanine Transaminase, BUN: Blood Urea Nitrogen, MIS: Malnutrition Inflammation Score, K-BDI: Korean-Beck Depression Inventory.

### 3.2 Outcomes

Potassium levels did not significantly differ between the groups (Table 2), no trend was observed (Table 3). Proportion of patients with normal levels at baseline was different (Table 2), but post-hoc tests were not significant after correcting for multiple testing (p=0.13, p=0.33) (Supplementary Table 3). Regression analysis did not result in any significant terms (Table 3).

**Table 2.**
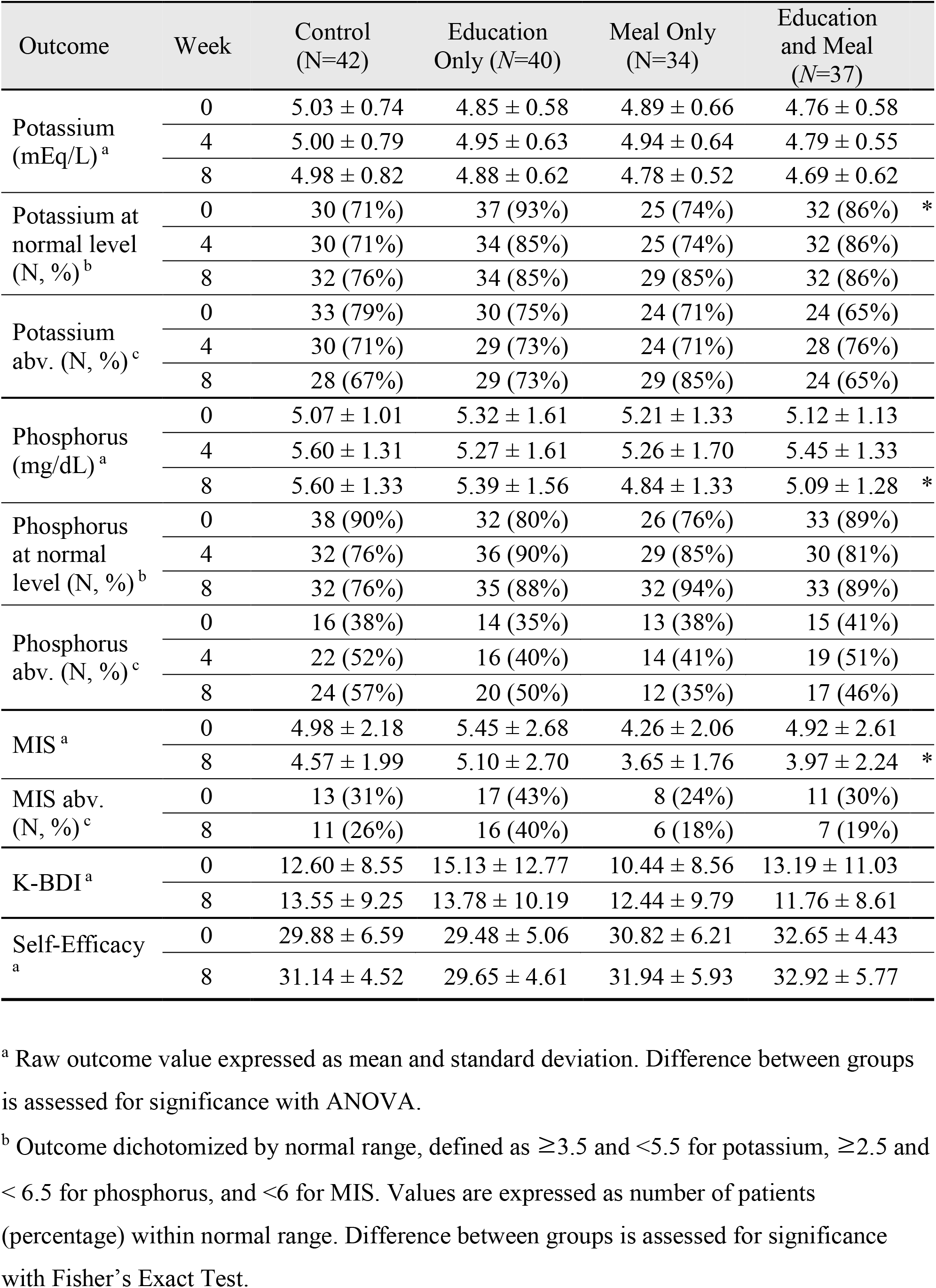
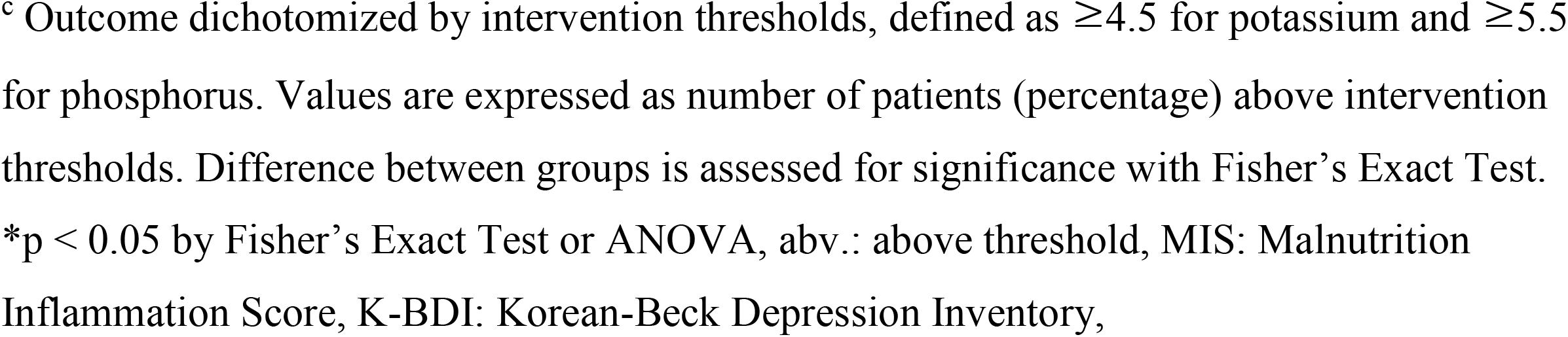
Analysis of differences in outcome by intervention group.

| Outcome | Week | Control<br>(N=42) | Education<br>Only (N=40) | Meal Only<br>(N=34) | Education<br>and Meal<br>(N=37) |
| --- | --- | --- | --- | --- | --- |
| Potassium<br>(mEq/L) <sup>a</sup> | 0 | 5.03 ± 0.74 | 4.85 ± 0.58 | 4.89 ± 0.66 | 4.76 ± 0.58 |
|  | 4 | 5.00 ± 0.79 | 4.95 ± 0.63 | 4.94 ± 0.64 | 4.79 ± 0.55 |
|  | 8 | 4.98 ± 0.82 | 4.88 ± 0.62 | 4.78 ± 0.52 | 4.69 ± 0.62 |
| Potassium at<br>normal level<br>(N, %) <sup>b</sup> | 0 | 30 (71%) | 37 (93%) | 25 (74%) | 32 (86%) * |
|  | 4 | 30 (71%) | 34 (85%) | 25 (74%) | 32 (86%) |
|  | 8 | 32 (76%) | 34 (85%) | 29 (85%) | 32 (86%) |
| Potassium<br>abv. (N, %) <sup>c</sup> | 0 | 33 (79%) | 30 (75%) | 24 (71%) | 24 (65%) |
|  | 4 | 30 (71%) | 29 (73%) | 24 (71%) | 28 (76%) |
|  | 8 | 28 (67%) | 29 (73%) | 29 (85%) | 24 (65%) |
| Phosphorus<br>(mg/dL) <sup>a</sup> | 0 | 5.07 ± 1.01 | 5.32 ± 1.61 | 5.21 ± 1.33 | 5.12 ± 1.13 |
|  | 4 | 5.60 ± 1.31 | 5.27 ± 1.61 | 5.26 ± 1.70 | 5.45 ± 1.33 |
|  | 8 | 5.60 ± 1.33 | 5.39 ± 1.56 | 4.84 ± 1.33 | 5.09 ± 1.28 * |
| Phosphorus<br>at normal<br>level (N, %) <sup>b</sup> | 0 | 38 (90%) | 32 (80%) | 26 (76%) | 33 (89%) |
|  | 4 | 32 (76%) | 36 (90%) | 29 (85%) | 30 (81%) |
|  | 8 | 32 (76%) | 35 (88%) | 32 (94%) | 33 (89%) |
| Phosphorus<br>abv. (N, %) <sup>c</sup> | 0 | 16 (38%) | 14 (35%) | 13 (38%) | 15 (41%) |
|  | 4 | 22 (52%) | 16 (40%) | 14 (41%) | 19 (51%) |
|  | 8 | 24 (57%) | 20 (50%) | 12 (35%) | 17 (46%) |
| MIS <sup>a</sup> | 0 | 4.98 ± 2.18 | 5.45 ± 2.68 | 4.26 ± 2.06 | 4.92 ± 2.61 |
|  | 8 | 4.57 ± 1.99 | 5.10 ± 2.70 | 3.65 ± 1.76 | 3.97 ± 2.24 * |
| MIS abv.<br>(N, %) <sup>c</sup> | 0 | 13 (31%) | 17 (43%) | 8 (24%) | 11 (30%) |
|  | 8 | 11 (26%) | 16 (40%) | 6 (18%) | 7 (19%) |
| K-BDI <sup>a</sup> | 0 | 12.60 ± 8.55 | 15.13 ± 12.77 | 10.44 ± 8.56 | 13.19 ± 11.03 |
|  | 8 | 13.55 ± 9.25 | 13.78 ± 10.19 | 12.44 ± 9.79 | 11.76 ± 8.61 |
| Self-Efficacy<br><sup>a</sup> | 0 | 29.88 ± 6.59 | 29.48 ± 5.06 | 30.82 ± 6.21 | 32.65 ± 4.43 |
|  | 8 | 31.14 ± 4.52 | 29.65 ± 4.61 | 31.94 ± 5.93 | 32.92 ± 5.77 |

<sup>a</sup> Raw outcome value expressed as mean and standard deviation. Difference between groups is assessed for significance with ANOVA.
<sup>b</sup> Outcome dichotomized by normal range, defined as $\geq 3.5$ and $< 5.5$ for potassium, $\geq 2.5$ and $< 6.5$ for phosphorus, and $< 6$ for MIS. Values are expressed as number of patients (percentage) within normal range. Difference between groups is assessed for significance with Fisher's Exact Test.
<sup>c</sup> Outcome dichotomized by intervention thresholds, defined as $\geq 4.5$ for potassium and $\geq 5.5$ for phosphorus. Values are expressed as number of patients (percentage) above intervention thresholds. Difference between groups is assessed for significance with Fisher's Exact Test. \* $p < 0.05$ by Fisher's Exact Test or ANOVA, abv.: above threshold, MIS: Malnutrition Inflammation Score, K-BDI: Korean-Beck Depression Inventory,

**Table 3.** Estimates of treatment effect.

| Outcome | Week | Education | Meal | Education and Meal |
| --- | --- | --- | --- | --- |
| *Change in mean outcome value<sup>a</sup>* |  |  |  |  |
| Potassium (mEq/L) | 4 | 0.06 [-0.17, 0.29] | 0.02 [-0.22, 0.26] | -0.04 [-0.45, 0.49] |
|  | 8 | -0.01 [-0.25, 0.24] | -0.13 [-0.38, 0.12] | -0.14 [-0.63, 0.37] |
| Phosphorus (mg/dL) | 4 | -0.44 [-1.04, 0.16] | -0.40 [-1.03, 0.23] | -0.17 [-1.64, 0.84] |
|  | 8 | -0.30 [-0.87, 0.27] | -0.81 [-1.40, -0.22]* | -0.52 [-1.98, 0.36] |
| MIS | 8 | 0.18 [-0.40, 0.76] | -0.40 [-1.01, 0.21] | -0.56 [-1.59, 0.80] |
| K-BDI | 8 | -1.41 [-4.34, 1.51] | 0.29 [-2.76, 3.34] | -2.18 [-5.73, 6.31] |
| Self-Efficacy | 8 | -1.34 [-3.41, 0.73] | 0.44 [-1.73, 2.61] | 0.72 [-3.84, 4.72] |
| *Odds ratio of outcome being in normal range<sup>b</sup>* |  |  |  |  |
| Potassium | 4 | 1.10 [0.93, 1.29] | 0.99 [0.83, 1.17] | 1.01 [0.79, 1.28] |
|  | 8 | 1.07 [0.91, 1.26] | 1.08 [0.91, 1.28] | 0.93 [0.73, 1.18] |
| Phosphorus | 4 | 1.17 [0.99, 1.37] | 1.11 [0.93, 1.31] | 0.82 [0.65, 1.04] |
|  | 8 | 1.14 [0.98, 1.31] | 1.21 [1.04, 1.40]* | 0.83 [0.67, 1.03] |
| MIS | 8 | 0.92 [0.80, 1.06] | 1.00 [0.86, 1.16] | 1.16 [0.94, 1.42] |

<sup>a</sup> Week 4 and 8 treatment effect [95% confidence interval] estimated as the mean outcome of intervention group minus the mean outcome of no intervention group. Estimate and 95% confidence intervals are derived from ordinary linear regression models controlling for baseline value.
<sup>b</sup> Week 4 and 8 treatment effect [95% confidence interval] estimated as the odds ratio of the intervention group having an outcome in normal range, compared to the control group. Normal ranges are defined as $\geq 3.5$ and $< 5.5$ for potassium, $\geq 2.5$ and $< 6.5$ for phosphorus, and $< 6$ for MIS. Log odds ratio and 95% confidence intervals are derived from logistic regression controlling for baseline value, then calculated as odds ratio.
\*p-value $< 0.05$ by ANOVA, MIS: Malnutrition Inflammation Score, K-BDI: Korean Beck Depression Inventory

Phosphorus levels decreased significantly at week 8 in the meal group (estimated change = −0.81 mEq/L, 95% C.I.=[-1.40, −0.22]) (Table 3). The control group increasing slope, which was not observed in other groups (Figure 3). Logistic model showed similar results, where meal increased the likelihood of having normal levels at week 8 (Odds ratio=1.21,95% C.I.=[1.04, 1.40]).

**Figure 3.**
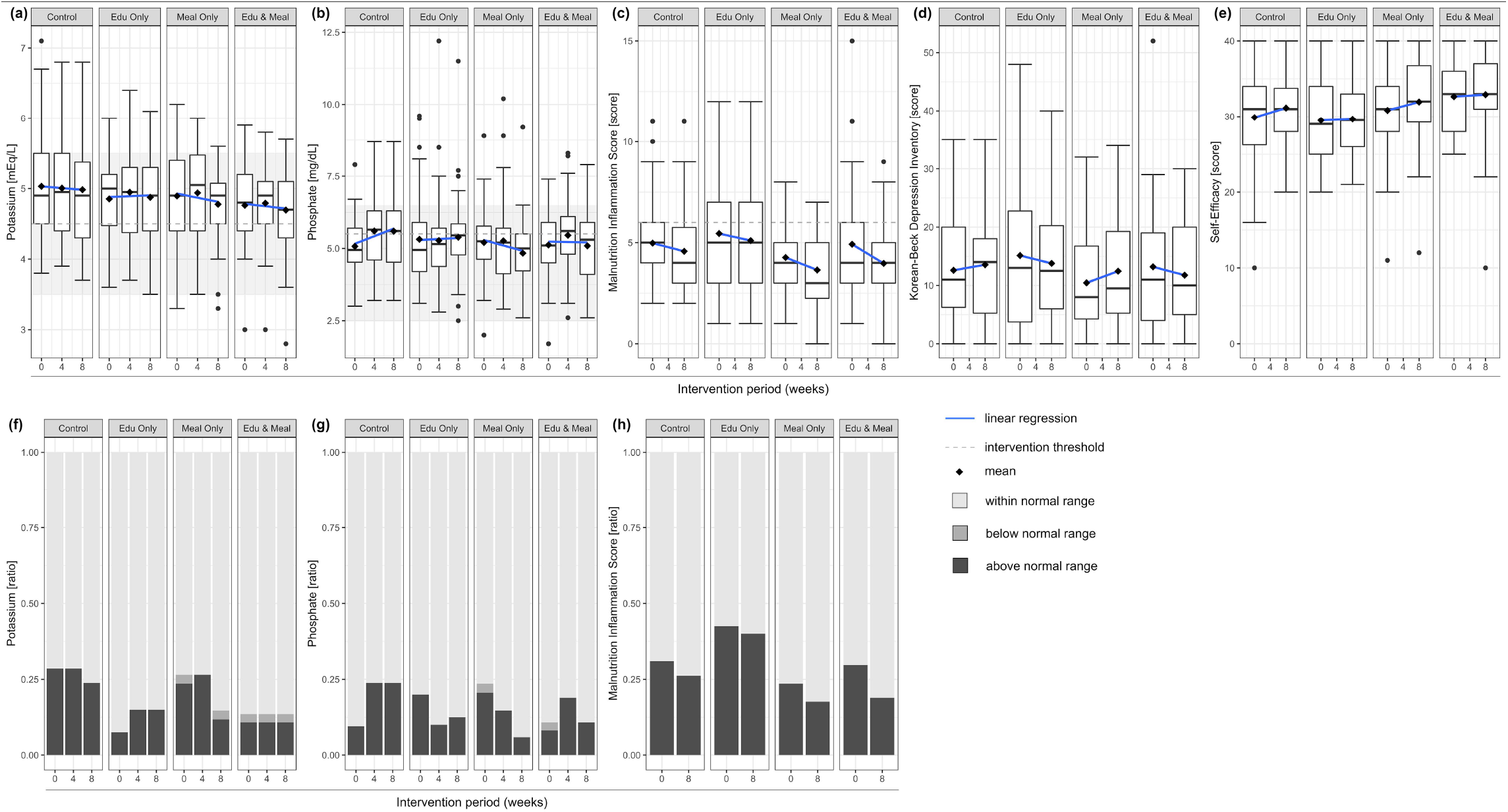
Outcome change of each intervention group by intervention period. (a-e) Distribution of outcome values are expressed as box-and-whisker plot. Mean values and linear regression lines have been plotted over in rhombus and solid line, respectively. Normal ranges of potassium and phosphate are shaded in the background. Intervention thresholds are drawn as horizontal dashed line, where applicable. (f-h) Ratio of subjects categorized by below, within, and above the normal ranges of each outcome. Normal ranges are defined as ≥3.5 mEq/L and <5.5 mEq/L for potassium, ≥2.5 mg/dL and < 6.5 mg/dL for phosphorus, and < 6 for Malnutrition Inflammation Score. Edu: Education.

MIS significantly differed between the groups at week 8 (Table 2); meal and education group had a significantly lower score compared to the control group (adjusted p=0.005) (Supplementary Table 3). There was no significant main or interaction effect of the interventions (Table 3). KBDI and Self-Efficacy did not significantly differ between groups (Table 2) or change with intervention (Table 3). However, education groups had a decreased score, while control and meal only groups had increased scores (Table 2, Figure 3d).

Meal compliance was high (88% meals consumed), and education satisfaction was above average (4.08 ± 0.71 points). Both scores did not significantly differ between the groups and did not have significant correlation with outcomes (Supplementary 22 patients who did not provide their phone numbers were not identifiable, thus excluded from analysis.).

### 3.3 Qualitative Lessons Learned

Dissatisfaction with the meal was a recurring issue. Example complaints were that there was too much rice, the seasonings were too strong, the dishes were not fresh, and there was too little protein variety. Concerns about the nutrient content was also an issue, as the meals included ingredients that patients had learned to avoid. Information leaflets describing the total nutritional content and its alignment with recommendations were not enough to relieve some patients. Some patients adjusted by skipping the side dish of issue, while others withdrew from the study completely.

The logistic process of meals was longer than expected. Placing the order, manufacturing the meals, and delivering the meal boxes to the patient’s homes took over four working days. Since the orders were placed on a weekly schedule, this could result in a total delay of up to 10 days in reflecting the patient’s latest lab values. As such, it was possible that the delivered meal did not correctly reflect the patient’s latest lab values at the time of consumption. Additionally, interruptions such as hospitalization, kidney transplant, unexpected travel days, and changes in recipient address resulted in several meals that were wasted.

## Discussion

Both meal and education did not significantly change potassium levels. Patients receiving education were more likely to have normal levels at baseline compared to their counterparts. However, potassium assessed as continuous variables did not significantly differ between the groups therefore this difference was not considered a severe violation of randomization.

On the other hand, patients receiving meals had a significantly lower phosphorus levels at week 8, compared to their counterparts. The control group had an increasing trend in phosphorus levels (0.26 per 4 weeks), which seems to be suppressed in groups receiving the meal intervention. Odds of being in normal range also increased, showing that phosphorus levels were not reduced inordinately. Such finding is meaningful, considering that both high and low serum phosphorus levels are associated with mortality in HD patients [26]. The increase in phosphorus levels of the control group is difficult to explain. In previous research, seasonal variations of phosphorus levels in HD patients were either insignificant or higher in the summer [27-30]. The current study spanned from October to January, so this cannot explain the increasing trend. A potential explanation is that South Koreans tend to consume more phosphate-rich food such as meat and fish in the winter [31]. Unfortunately, a detailed study on phosphorus level variations of Korean HD patients could not be found.

Education did not show a significant effect on phosphorus levels, but the direction of change was consistently negative at both weeks 4 (β = −0.44) and 8 (β = −0.30). Previous studies found phosphate-specific diet education to significantly reduce serum phosphorus [32, 33]. In this study, phosphate education was provided in conjunction with materials on other topics such as protein. The importance of dietary phosphate may have not been as strongly emphasized, thus was not as effective as phosphate-specific education. Interestingly, the meal and education group did not show significant changes. Receiving both interventions could have influenced unobserved dietary behavior, such as consuming more phosphorus-rich protein. Nevertheless, further study is needed to find a plausible explanation.

Overall, the intervention resulted in desirable changes in controlling serum phosphorus, but not for serum potassium. The disparity could reflect how patients are better at controlling dietary potassium than phosphate, perhaps driven by differences in food preference, emphasis in nutritional education, or perceived associated risks. The fact that some patients refused meals out of concern for excess potassium, but no concerns were raised about phosphate, further suggests such possibility. Likewise, studies have found that HD patients are the least compliant with dietary restriction of phosphate, compared to potassium and sodium [34]. Some studies suggest that limiting dietary potassium in HD patients is a standard practice that lacks scientific evidence of reducing hyperkalemia or mortality [35, 36]. Results of this study further supports the idea that dietary control of potassium is not well reflected in serum levels.

MIS significantly decreased, thus improved, in groups receiving meal intervention at week 8, compared to their counterparts. Although not at significance, MIS decreased and self-efficacy increased in all intervention groups. Participation in the study and completing health-related surveys could have increased general awareness in nutrition and self-care. None of the measures had significant negative changes from the intervention, while bringing desirable changes to phosphorus control. Since measures of malnutrition, depression, and self-efficacy may not change over a short time, longer follow-up time may be needed to observe significant changes.

### Limitations and Future work

The authors recognize many limitations of this study. Data was not collected for potential factors of influence, such as survey on other meals consumed, usual dietary habits, and details on binder prescription. Meal variety was only limited to two types and type B comprised 96% of the meal delivered. Intervention efficacy could be increased by expanding variety to allow tailoring at increased granularity, replacing more than one daily meal, and targeting a subpopulation that experience more difficulty with meal preparation. The tailoring algorithm was solely based on clinical values, whereas considering for patient’s social and behavioral characteristics could have been more effective. Future work should develop the algorithm to include more social, behavioral, or cultural factors.

Promoting the App usage was an important goal of the grant funding this study, but adoption rate was low; 53% of the study participants was registered, and only 27% had more than one login day during the trial. The intervention algorithm was intended to function on the platform; thus, patients should have received the educational materials and surveys on the App, which was not realized. Although not fully streamlined, the algorithm was executed by an intermediary program that automatically processed participant’s data. Education materials in the App could have had an influence on the outcomes, but its effect was considered insignificant, considering that randomization was executed independently, no difference was found between the groups, and overall usage rate was low.

The authors plan to improve and implement this adaptive intervention algorithm into the existing platform. The algorithm can access and process patient data on the platform, streamlining the process of generating meal orders and providing educational materials. This will minimize delay and enable incorporation of various health data into the algorithm, ensuring its reliability, scalability, and diversity.

## Practical Application

The algorithm in this study can be further developed and incorporated into any health information systems to provide automatic tailored intervention for hemodialysis patients. Specifically, serum phosphorus levels can be stabilized via targeting patients with levels above the threshold of 5.5 mg/dL. Providing low-potassium and low-phosphate meals can be an effective means to control serum phosphorus, with no significant negative effects on other nutritional markers.

## Supporting information

CONSORT checklist

## Data Availability

All data produced in the present study are available upon reasonable request to the authors

## Acknowledgements

The Chronic Kidney Disease Common Data Model Special Interest Group, a member of the Korean Society of Nephrology, have long been facilitators of the nation-wide multi-center cohort of this study. Eatmapl Inc. contributed to the study by manufacturing, storing, and delivering specialized meals for hemodialysis patients. AvChain, Inc. contributed mobile applications for the decentralized clinical trial.

