## Supplementary material for "Adaptive nutrition intervention stabilizes serum phosphorus levels in hemodialysis patients: a multi-center decentralized clinical trial using real-world data": CONSORT checklist

### Reporting checklist for randomised trial.

Based on the CONSORT guidelines.

#### Instructions to authors

Complete this checklist by entering the page numbers from your manuscript where readers will find each of the items listed below.

Your article may not currently address all the items on the checklist. Please modify your text to include the missing information. If you are certain that an item does not apply, please write "n/a" and provide a short explanation.

Upload your completed checklist as an extra file when you submit to a journal.

In your methods section, say that you used the CONSORTreporting guidelines, and cite them as:

Schulz KF, Altman DG, Moher D, for the CONSORT Group. CONSORT 2010 Statement: updated guidelines for reporting parallel group randomised trials

|  |  | Reporting Item | Page Number |
| --- | --- | --- | --- |
| **Title and Abstract** |  |  |  |
| Title | [#1a](https://www.goodreports.org/reporting-checklists/consort/info/#1a) | Identification as a randomized trial in the title. | n/a, in separate file |
| Abstract | [#1b](https://www.goodreports.org/reporting-checklists/consort/info/#1b) | Structured summary of trial design, methods, results, and conclusions | 1 |
| **Introduction** |  |  |  |
| Background and objectives | [#2a](https://www.goodreports.org/reporting-checklists/consort/info/#2a) | Scientific background and explanation of rationale | 2 |
| Background and objectives | [#2b](https://www.goodreports.org/reporting-checklists/consort/info/#2b) | Specific objectives or hypothesis | 2 |
| **Methods** |  |  |  |
| Trial design | [#3a](https://www.goodreports.org/reporting-checklists/consort/info/#3a) | Description of trial design (such as parallel, factorial) including allocation ratio. | 3 |
| Trial design | [#3b](https://www.goodreports.org/reporting-checklists/consort/info/#3b) | Important changes to methods after trial commencement (such as eligibility criteria), with reasons | 3 |
| Participants | [#4a](https://www.goodreports.org/reporting-checklists/consort/info/#4a) | Eligibility criteria for participants | 3 |
| Participants | [#4b](https://www.goodreports.org/reporting-checklists/consort/info/#4b) | Settings and locations where the data were collected | 3 |
| Interventions | [#5](https://www.goodreports.org/reporting-checklists/consort/info/#5) | The experimental and control interventions for each group with sufficient details to allow replication, including how and when they were actually administered | 4 |
| Outcomes | [#6a](https://www.goodreports.org/reporting-checklists/consort/info/#6a) | Completely defined prespecified primary and secondary outcome measures, including how and when they were assessed | 5 |
| Sample size | [#7a](https://www.goodreports.org/reporting-checklists/consort/info/#7a) | How sample size was determined. | 3 |
| Sample size | [#7b](https://www.goodreports.org/reporting-checklists/consort/info/#7b) | When applicable, explanation of any interim analyses and stopping guidelines | 3 |
| Randomization - Sequence generation | [#8a](https://www.goodreports.org/reporting-checklists/consort/info/#8a) | Method used to generate the random allocation sequence. |  |
| 3 |  |  |  |
| Randomization - Sequence generation | [#8b](https://www.goodreports.org/reporting-checklists/consort/info/#8b) | Type of randomization; details of any restriction (such as blocking and block size) |  |
| 3 |  |  |  |
| Randomization - Allocation concealment mechanism | [#9](https://www.goodreports.org/reporting-checklists/consort/info/#9) | Mechanism used to implement the random allocation sequence (such as sequentially numbered containers), describing any steps taken to conceal the sequence until interventions were assigned | 3 |
| Randomization - Implementation | [#10](https://www.goodreports.org/reporting-checklists/consort/info/#10) | Who generated the allocation sequence, who enrolled participants, and who assigned participants to interventions | 3 |
| Blinding | [#11a](https://www.goodreports.org/reporting-checklists/consort/info/#11a) | If done, who was blinded after assignment to interventions (for example, participants, care providers, those assessing outcomes) and how. | 3 |
| Blinding | [#11b](https://www.goodreports.org/reporting-checklists/consort/info/#11b) | If relevant, description of the similarity of interventions | 3 |
| Statistical methods | [#12a](https://www.goodreports.org/reporting-checklists/consort/info/#12a) | Statistical methods used to compare groups for primary and secondary outcomes | 5 |
| Statistical methods | [#12b](https://www.goodreports.org/reporting-checklists/consort/info/#12b) | Methods for additional analyses, such as subgroup analyses and adjusted analyses | 5 |
| Outcomes | [#6b](https://www.goodreports.org/reporting-checklists/consort/info/#6b) | Any changes to trial outcomes after the trial commenced, with reasons | 5 |
| **Results** |  |  |  |
| Participant flow diagram (strongly recommended) | [#13a](https://www.goodreports.org/reporting-checklists/consort/info/#13a) | For each group, the numbers of participants who were randomly assigned, received intended treatment, and were analysed for the primary outcome | 16 |
| Participant flow | [#13b](https://www.goodreports.org/reporting-checklists/consort/info/#13b) | For each group, losses and exclusions after randomization, together with reason | 16 |
| Recruitment | [#14a](https://www.goodreports.org/reporting-checklists/consort/info/#14a) | Dates defining the periods of recruitment and follow-up | 3 |
| Recruitment | [#14b](https://www.goodreports.org/reporting-checklists/consort/info/#14b) | Why the trial ended or was stopped | 3 |
| Baseline data | [#15](https://www.goodreports.org/reporting-checklists/consort/info/#15) | A table showing baseline demographic and clinical characteristics for each group | 19 |
| Numbers analysed | [#16](https://www.goodreports.org/reporting-checklists/consort/info/#16) | For each group, number of participants (denominator) included in each analysis and whether the analysis was by original assigned groups | 19 |
| Outcomes and estimation | [#17a](https://www.goodreports.org/reporting-checklists/consort/info/#17a) | For each primary and secondary outcome, results for each group, and the estimated effect size and its precision (such as 95% confidence interval) | 23 |
| Outcomes and estimation | [#17b](https://www.goodreports.org/reporting-checklists/consort/info/#17b) | For binary outcomes, presentation of both absolute and relative effect sizes is recommended | 23 |
| Ancillary analyses | [#18](https://www.goodreports.org/reporting-checklists/consort/info/#18) | Results of any other analyses performed, including subgroup analyses and adjusted analyses, distinguishing pre-specified from exploratory | n/a, none performed |
| Harms | [#19](https://www.goodreports.org/reporting-checklists/consort/info/#19) | All important harms or unintended effects in each group (For specific guidance see CONSORT for harms) | n/a, no harm found |
| **Discussion** |  |  |  |
| Limitations | [#20](https://www.goodreports.org/reporting-checklists/consort/info/#20) | Trial limitations, addressing sources of potential bias, imprecision, and, if relevant, multiplicity of analyses | 10 |
| Interpretation | [#22](https://www.goodreports.org/reporting-checklists/consort/info/#22) | Interpretation consistent with results, balancing benefits and harms, and considering other relevant evidence | 8 |
| Registration | [#23](https://www.goodreports.org/reporting-checklists/consort/info/#23) | Registration number and name of trial registry | n/a, in separate file |
| Generalisability | [#21](https://www.goodreports.org/reporting-checklists/consort/info/#21) | Generalisability (external validity, applicability) of the trial findings | 10 |
| **Other information** |  |  |  |
| Interpretation | [#22](https://www.goodreports.org/reporting-checklists/consort/info/#22) | Interpretation consistent with results, balancing benefits and harms, and considering other relevant evidence | 8 |
| Registration | [#23](https://www.goodreports.org/reporting-checklists/consort/info/#23) | Registration number and name of trial registry | n/a, in separate file |
| Protocol | [#24](https://www.goodreports.org/reporting-checklists/consort/info/#24) | Where the full trial protocol can be accessed, if available | n/a, in separate file |
| Funding | [#25](https://www.goodreports.org/reporting-checklists/consort/info/#25) | Sources of funding and other support (such as supply of drugs), role of funders | n/a, in separate file |

Notes:

- 1a: n/a, in separate file
- 18: n/a, none performed
- 19: n/a, no harm found
- 23: n/a, in separate file
- 24: n/a, in separate file
- 25: n/a, in separate file The CONSORT checklist is distributed under the terms of the Creative Commons Attribution License CC-BY. This checklist was completed on 20. February 2023 using <https://www.goodreports.org/>, a tool made by the [EQUATOR Network](https://www.equator-network.org) in collaboration with [Penelope.ai](https://www.penelope.ai)
